# PFAS Exposure During Pregnancy Is Associated with Shorter Gestational Age at Birth and with Growth Trajectories During the First 2 Years of Life

**DOI:** 10.64898/2026.10.20.26366022

**Authors:** Kyle A. Campbell, Clark R. Sims, Priyadarshni Patel, Lauren A. Havens, Andrew J. Morris, Kartik Shankar, Kevin J. Pearson, Aline Andres, Todd M. Everson

**Affiliations:** Gangarosa Department of Environmental Health, Rollins School of Public Health at Emory University, Atlanta, Georgia, United States; Arkansas Children’s Nutrition Center, Little Rock, Arkansas, United States; Department of Pediatrics, University of Arkansas for Medical Sciences, Little Rock, Arkansas, United States; Central Arkansas Veterans Affairs Healthcare System and University of Arkansas for Medical Sciences, Little Rock, Arkansas, United States; Responsive Agricultural Food Systems Research Unit, Plains Area, USDA Agricultural Research Service, College Station, Texas, United States; Department of Nutrition, Texas A&M University, College Station, Texas, United States; Department of Pharmacology & Nutritional Sciences, University of Kentucky College of Medicine, Lexington, Kentucky, United States; Department of Epidemiology, Rollins School of Public Health at Emory University, Atlanta, Georgia, United States

**Keywords:** Per- and polyfluoroalkyl substances, PFAS, pregnancy, infant growth, fat mass, quantile g-computation

## Abstract

**Introduction:** Prenatal exposure to per- and polyfluoroalkyl substances (PFAS) may influence birth outcomes and postnatal growth and adiposity, yet prospective evidence is limited. We evaluated sex-specific associations of first trimester maternal PFAS exposure with birth outcomes and children’s growth and body composition through age two.

**Methods:** Participants were enrolled in the Glowing prospective cohort study (2010-2014, NCT01131117). Levels of six PFAS (PFOS, PFOA, PFBS, PFHxS, PFNA, PFHxA) in maternal serum at 10-12 weeks’ gestation were analyzed. Five participants were excluded due to unsuccessful PFAS characterization. We tested associations between PFAS exposure and gestational age, birth weight, birth length, and 11 repeated measures of weight, length, fat mass, and fat-free mass (FFM) during the first two years of life with linear regression, SuperImposition by Translation And Rotation growth curve size and intensity, and quantile g-computation.

**Results:** Among 217 complete-case dyads, higher first trimester PFAS concentrations were associated with shorter gestation (PFOS and PFHxS; e.g., PFOS β = −0.198 weeks per doubling, 95% CI −0.368 to −0.028). PFHxS was associated with lower birth weight (β = −0.048 kg, 95% CI −0.093 to −0.002). Among males only, PFOS, PFHxS, PFOA, and PFNA were associated with shorter gestation and PFNA with shorter birth length. For postnatal growth, PFBS was associated with lower weight and fat-free mass growth intensity. PFHxS was associated with smaller length and fat-free mass among females. The joint six-PFAS mixture was not associated with any outcome. Associations were robust to adjustment for maternal albumin and estimated glomerular filtration rate.

**Conclusion:** In a Central Arkansas birth cohort, first trimester maternal PFAS concentrations, particularly PFHxS, were individually associated with shorter gestation and reduced early growth and fat-free mass accrual, with more sex-specific male associations among birth outcomes and more female associations in longitudinal postnatal growth through age two.

**Graphical Abstract:** 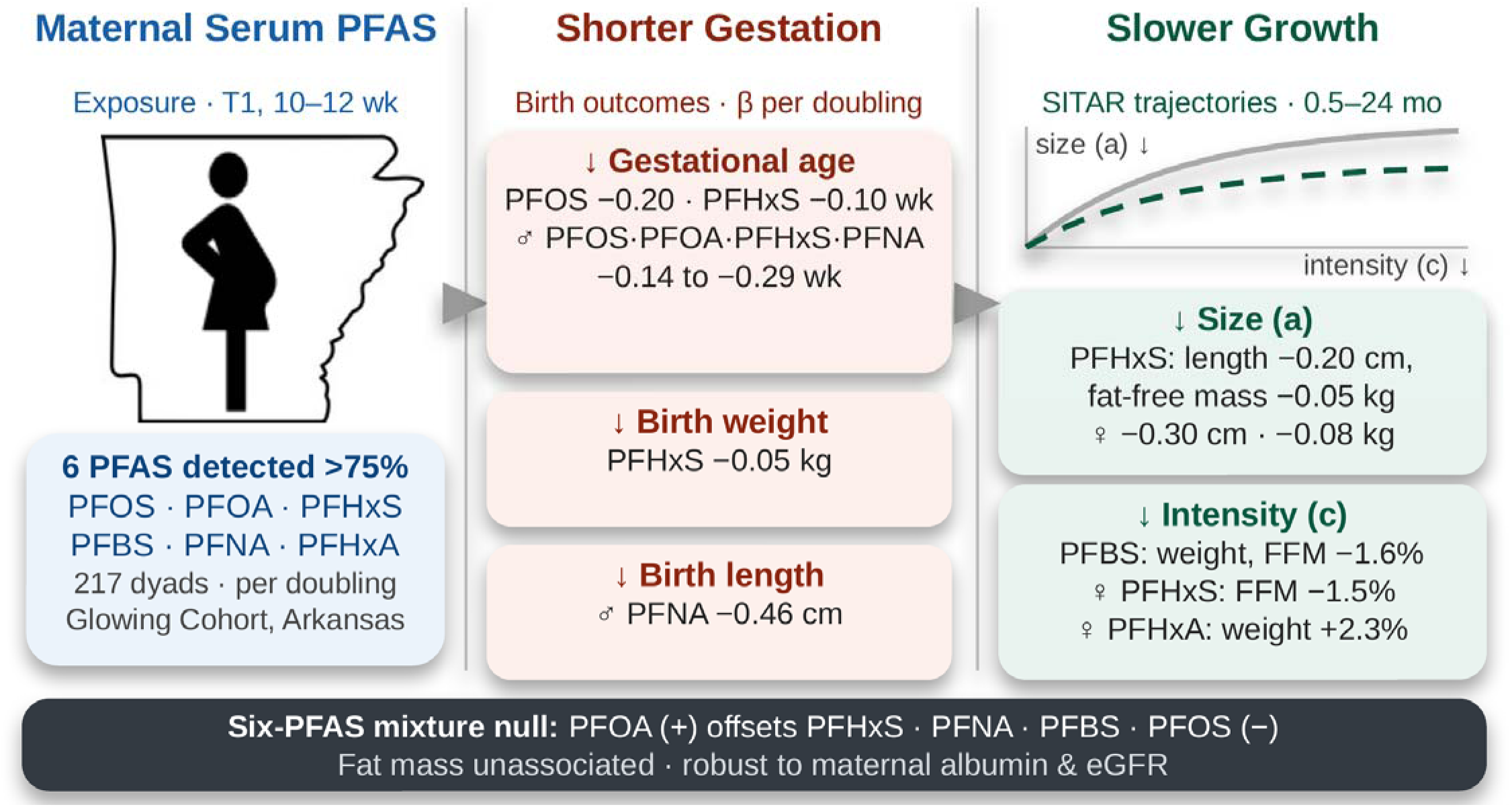

## Introduction

Environmental exposures during the perinatal period can have profound and long-term impacts on health^1^. Per- and polyfluoroalkyl substances (PFAS), a family of synthetic chemicals characterized by strong carbon-fluorine bonds, are of particular concern given their ubiquity and persistence^2^. Over 98% of Americans have detectable concentrations of PFAS in serum^3^. Although some legacy PFAS have been phased out, their environmental stability and slow elimination from the body result in persistent and widespread human exposure^4^. In addition, numerous other PFAS are still being developed and produced for consumer products and industrial applications. Because PFAS contamination can vary substantially across regions, geographic location is a major determinant of PFAS exposure^5^. To our knowledge, no published study has evaluated prenatal PFAS exposure and infant health outcomes in this region and exposure setting.

Prenatally, PFAS are known to contribute to growth restriction and influence the risk of several chronic diseases later in life. Prenatal exposure to PFAS has been associated with smaller birth size in numerous studies^6–9^ and preterm birth or shorter gestation^6,10–12^, as well as several children’s health outcomes including risk of obesity^13–15^, and, in later childhood, greater adiposity and body mass index (BMI) or weight-for-length z-score^16–18^. While associations with smaller birth size have been relatively consistent, effects on postnatal growth and adiposity remain less clear due to a scarcity of longitudinal cohorts with repeated body composition measures^16^. Some studies of children’s growth found no sex differences in associations between prenatal PFAS exposures and children’s BMI z-scores, fat mass, or waist circumference^19,20^. Most prior studies have also relied on weight and BMI or other proxies of adiposity rather than direct measures of fat and fat-free mass, have characterized exposure to individual PFAS rather than the correlated mixture to which fetuses are exposed, and have rarely evaluated whether pregnancy hemodynamics might confound the observed associations.

There is growing evidence that PFAS-associated outcomes may differ by fetal sex, though the direction has varied across cohorts and outcomes. Data from the Environmental Influences on Child Health Outcomes cohorts showed that PFAS-associated birth outcomes differed by sex, where PFAS-associated reductions in growth were strongest in female newborns^21^ and a study of NHANES 2013-2016 participants found that PFAS were associated with poor liver biomarker profiles in females but not in males^22^. One hypothesis holds that the male placenta prioritizes growth over adaptation to an adverse intrauterine environment, leaving males more vulnerable to insults at birth, whereas the female placenta adapts to buffer early stress at the possible cost of later developmental effects^23^. Consistent with a biological basis for such dimorphism, many effects of PFAS are mediated through peroxisome proliferator-activated receptors, which are known to act in a sex-specific manner^24,25^; however, it is unclear if PFAS play a role.

This study evaluated associations of first trimester maternal PFAS exposure with children’s growth and body composition across the first two years of life in a cohort residing in Central Arkansas. We characterized growth using quantitative fat and fat-free mass in addition to weight and length, summarized repeated measures of detailed follow-up with trajectory modeling, and examined individual PFAS, their joint mixture effect, and the potential for confounding by maternal hemodynamics. We hypothesized that higher prenatal PFAS exposure would be associated with early growth restriction, including reduced gestational age, birth length, or birth weight, followed by differences in body composition during the first two years of life, and that some of these relationships would be sex-specific.

## Methods

### Cohort

Participants were enrolled in the Glowing longitudinal study between 2010 and 2014 (www.clinicaltrials.gov, ID # NCT01131117). The original cohort consisted of 300 dyads (pregnant women and offspring) who were enrolled prior to gestation week 10. All participants were of second parity, pregnant with a single fetus, non-smokers, reported no alcohol use and were older than 21 years of age. Exclusion criteria included pre-existing medical conditions, sexually transmitted infections, medical complications or medications during pregnancy known to influence fetal growth (including gestational diabetes, thyroid disease, pre-eclampsia, etc.) and conceptions aided with fertility treatment. Infants included in the analyses were full-term (≥ 37 weeks’ gestation) and had no medical conditions and required no medications known to influence growth or development of the fetus. A total of 222 dyads completed the study at post-partum age 2 years (74%)^26^ and had samples available for PFAS evaluation. Five dyads did not have enough serum sample for the PFAS characterization. All participants provided informed consent prior to any study procedures. The protocol was approved by the Institutional Review Board of the University of Arkansas for Medical Sciences.

### Pregnant women self-reported variables and anthropometric measurements

Participants self-reported race, date of birth, education level, delivery method, child date of birth, child sex, birth weight, and birth length. Gestational age at birth was calculated from the estimated due date obtained by ultrasound. Anthropometric measures were obtained at each study visit using standardized methods. Maternal body weight was measured to the nearest 0.1 kg on a tared scale (Perspective Enterprises, Portage, MI, USA). Standing height was measured using a standard wall-mounted stadiometer to the nearest 0.1 cm (Tanita Corp., Tokyo, Japan) at enrollment. Body mass index was calculated (BMI: weight [kg]/height [m]²) for all participants.

### Per- and polyfluoroalkyl substances maternal serum quantification

Per- and polyfluoroalkyl substances (PFAS) were measured in maternal serum during the first study visit at 10-12 weeks of pregnancy following an overnight fast using UHPLC coupled electrospray ionization tandem mass spectrometry and adaptations of previous published methods^27–31^. The following PFASs were measured: perfluorobutanoic acid (PFBA), perfluorobutane sulfonate (PFBS), perfluoropentane sulfonic acid (PFPeS), perfluorohexane sulfonate (PFHxS), perfluoroheptanesulfonic acid (PFHpS), perfluorooctane sulfonic acid (PFOS), perfluorononanesulfonic acid (PFNS), perfluorodecanesulfonic acid (PFDS), perfluoro-n-pentanoic acid (PFPeA), perfluorohexanoic Acid (PFHxA), perfluoroheptanoic acid (PFHpA), perfluorooctanoic acid (PFOA), perfluorononanoic acid (PFNA), perfluorodecanoic acid (PFDA), perfluoroundecanoic acid (PFUnDA), perfluorododecanoic acid (PFDoA), perfluorotridecanoic acid (PFTrDA), perfluorotetradecanoic acid (PFTeDA), hexafluoropropylene oxide dimer acid (HFPO-DA), 4,8-dioxa-3H-perfluorononanoic acid (DONA), 9-chlorohexadecafluoro-3-oxanonane-1-sulfonic acid (9Cl-PF3ONS), 11-chloroeicosafluoro-3-oxaundecane-1-sulfonic acid (11Cl-PF3OUdS), perfluorooctanesulfonamide (PFOSA), n-methylperfluorooctanesulfonamidoacetic acid (N-MeFOSAA), n-ethyl perfluorooctane sulfonamido acetic acid (N-EtFOSAA), 4:2 fluorotelomer sulfonic acid (4:2 FTS), 6:2 fluorotelomer sulfonic acid (6:2 FTS), and 8:2 fluorotelomer sulfonic acid (8:2 FTS). Materials used as calibrators, controls, and mass labeled internal standards where available were purchased from Wellington Laboratories (Guelph, Ontario, Canada). The limit of detection (LOD) for all PFAS was 0.1 ng/ml and concentrations below the LOD were replaced with the LOD/√2^32^. Only PFOS, PFOA, PFBS, PFHxS, PFNA and PFHxA were detected in greater than 75% of the samples and served as the primary exposures of interest.

### Child growth and body composition

Child growth and body composition measures were obtained at 0.5, 1, 2, 3, 4, 5, 6, 9, 12, 18 and 24 months using standardized techniques. Weight was measured using a tared scale (SECA Corp, Ontario, Canada) to the closest 0.1 kg and length was measured using a length board with a sliding foot piece to the closest 0.1 cm (Perspective Enterprises). Infant fat and fat-free mass were obtained using quantitative nuclear magnetic resonance (EchoMRI-AH, Echo Medical Systems) to capture fat-free mass and fat mass measures as previously described^33^.

### Breastfeeding duration

Breast feeding duration was assessed via maternal self-report at each postnatal visit and was reported in weeks. If a child stopped breastfeeding between research study visits, the research assistant asked the participant’s caregiver for the exact date of breastfeeding cessation (defined as the cessation of nursing and of expressed human milk use).

### Statistical Analysis

Analyses were performed in R version 4.5.1. Maternal and infant characteristics were summarized as mean (SD) for continuous variables and count (%) for categorical variables. Maternal serum PFAS concentrations were summarized using geometric means and geometric standard deviations, with the proportion of samples above the limit of detection reported for each analyte. Analyses were restricted to the six PFAS detected in more than 75% of samples (PFOS, PFOA, PFBS, PFHxS, PFNA, and PFHxA). For all models, PFAS concentrations were log_2_-transformed so that regression coefficients corresponded to the difference in outcome per doubling of exposure.

Individual growth trajectories from 0.5 to 24 months were characterized using SuperImposition by Translation And Rotation (SITAR) mixed-effects models, fit separately by sex for weight, length, fat mass, and fat-free mass. Each model estimated subject-specific random effects for size (a) and intensity (c); a timing parameter (b) was not modeled due to convergence issues. The size parameter (a) reflects an individual’s overall magnitude on the growth curve (larger or smaller than average at a given age), while the intensity parameter (c) reflects the pace of growth, with higher values corresponding to an earlier, higher peak growth velocity. The size and intensity parameters were extracted for use as outcomes.

Associations between individual PFAS and each outcome were estimated using separate linear regression models in single congener PFAS models. For SITAR size and intensity parameters, models were adjusted for maternal BMI, maternal age, maternal education, and breastfeeding duration; birth-outcome models were adjusted for maternal BMI, maternal age, and maternal education. Self-reported race was not included as a covariate because the distribution was highly unbalanced (88% White) and the remaining participants comprised heterogeneous groups too small to support a stable or interpretable contrast. All models were fit in the pooled cohort, which included a sex covariate, and in separate sex-stratified models. Growth curve models were fit using the sitar R package, and mixture models used the qgcomp R package.

The joint effect of the PFAS mixture was estimated with quantile g-computation. Exposures were categorized into quartiles, and the mixture effect (ψ) represents the change in outcome for a simultaneous one-quartile increase in all six PFAS. These models used the same covariate adjustment as the single congener PFAS analyses and were fitted in the pooled cohort and in separate sex-stratified models. To characterize the directional contributions of individual PFAS to the joint effect, we extracted the positive and negative weights from each primary quantile g-computation fit. Each weight is the proportion of its same-signed partition attributable to a given PFAS, and the partition-specific sums (ψ+ and ψ−) indicate the extent to which opposing component effects offset one another within the overall ψ. Weights were visualized as diverging bar charts.

Because pregnancy is accompanied by plasma volume expansion and increased glomerular filtration, maternal serum PFAS concentrations may partly reflect maternal hemodynamics rather than exposure alone^34–36^. To evaluate this possibility, we performed sensitivity analyses repeating all single-congener and quantile g-computation models under three additional covariate sets on top of the primary adjustment factors: (i) first trimester serum albumin (g/dL); (ii) first trimester estimated glomerular filtration rate (eGFR, CKD-EPI^37^); and (iii) both markers simultaneously. Sensitivity models were fit for all birth and SITAR outcomes in the pooled cohort and in sex-stratified models, using the same quartile categorization (q = 4) as the primary analysis. Because these analyses were restricted to participants with complete hemodynamic data, sample sizes were reduced relative to the primary analysis: in the pooled cohort, n = 186 for albumin-adjusted, n = 185 for eGFR-adjusted, and n = 185 for jointly adjusted models, compared with n = 217 in the primary analysis (females: n = 77 for each sensitivity model versus n = 93; males: n = 109, 108, and 108 versus n = 124).

## Results

### Descriptives

A total of 217 complete-case mother–child Glowing study dyads from the Central Arkansas region with maternal serum PFAS characterization during the first study visit at 10-12 weeks gestation and postnatal growth data available were included in this analysis (**Table 1**). On average, participants were 29.5 years of age (SD 3.5), mostly white (88%) and with a college degree (65%). Their average BMI was 26 kg/m² (SD 4.3) and most delivered vaginally (65%). Participants’ newborns were born, on average, at 39.3 weeks of gestational age (SD 1.1), 3.5 kg (SD 0.5) in birth weight, and 51 cm (SD 2.6) in birth length. Slightly more than half of infants were male (57%), and infants were breastfed for an average of 6.7 months (SD 6.6). The standard deviation in breastfeeding duration was high because 43 women reported no breastfeeding at all.

**Table 1.** Glowing participants analytic sample (Central Arkansas, 2010-2014)

| Characteristic | N = 217<sup>1</sup> |
| --- | --- |
| Maternal age (y) | 29.46 (3.53) |
| Maternal BMI (kg/m<sup>2</sup>) | 25.93 (4.25) |
| Maternal education |  |
| College Degree | 141 (65%) |
| No College Degree | 76 (35%) |
| Maternal race |  |
| White | 190 (88%) |
| Other (collapsed<sup>2</sup>) | 25 (12%) |
| Not reported | 2 |
| Delivery method |  |
| C-Section | 77 (35%) |
| Vaginal | 140 (65%) |
| Child sex |  |
| Female | 93 (43%) |
| Male | 124 (57%) |
| Birth weight (kg) | 3.52 (0.49) |
| Birth length (cm) | 51.07 (2.61) |
| Gestational age (wk) | 39.28 (1.05) |
| Breastfeeding duration (mo) | 6.66 (6.56) |

<sup>1</sup> Mean (SD); n (%)
<sup>2</sup> Collapsed due to small cell size

The six PFAS detected in more than 75% of first trimester samples were PFOS (100%), PFOA (97%), PFHxS (93%), PFBS (92%), PFNA (90%), and PFHxA (86%, **Table 2**), with PFOS having the highest concentrations (geometric mean = 3.66 ng/ml) and PFBS the lowest (0.31 ng/ml). Because PFAS concentrations were log_2_-transformed for all analyses, single-congener PFAS regression coefficients correspond to the difference in outcome per doubling of exposure, and mixture effects (Ψ) correspond to a simultaneous one-quartile increase in all six PFAS.

**Table 2.** Exposure descriptive statistics for the analytic sample.

| PFAS | Geometric Mean (ng/mL) | Geometric SD | Number of observations above LOD | Percent above LOD (%) |
| --- | --- | --- | --- | --- |
| PFOS | 3.66 | 1.75 | 217 | 100 |
| PFOA | 1.02 | 2.34 | 210 | 96.8 |
| PFBS | 0.31 | 2.05 | 199 | 91.7 |
| PFHxS | 0.63 | 2.7 | 201 | 92.6 |
| PFNA | 0.43 | 2.26 | 196 | 90.3 |
| PFHxA | 0.44 | 2.42 | 187 | 86.2 |

### Associations between prenatal PFAS and anthropometric birth outcomes

In single PFAS congener models adjusted for maternal BMI, age, and education, higher maternal PFAS concentrations were associated with shorter gestation, including among males alone. In the whole cohort, each doubling of PFOS was associated with a shorter gestational age (β = −0.198 wk, 95% CI: −0.368 to −0.028, p = 0.024, **Figure 1**), as was PFHxS (β = −0.098 wk, 95% CI: −0.195 to −0.001, p = 0.048). A negative trend was also observed between PFOA and gestational age in the whole cohort (β = −0.105 wk, 95% CI: −0.218 to 0.008, p = 0.071). In males, gestational age was inversely associated with PFNA (β = −0.243 wk, 95% CI: −0.422 to −0.064, p = 0.009), PFOS (β = −0.289 wk, 95% CI: −0.524 to −0.054, p = 0.018), PFOA (β = −0.215 wk, 95% CI: −0.387 to −0.044, p = 0.015), and PFHxS (β = −0.144 wk, 95% CI: −0.284 to −0.005, p = 0.045).

**Figure 1.**
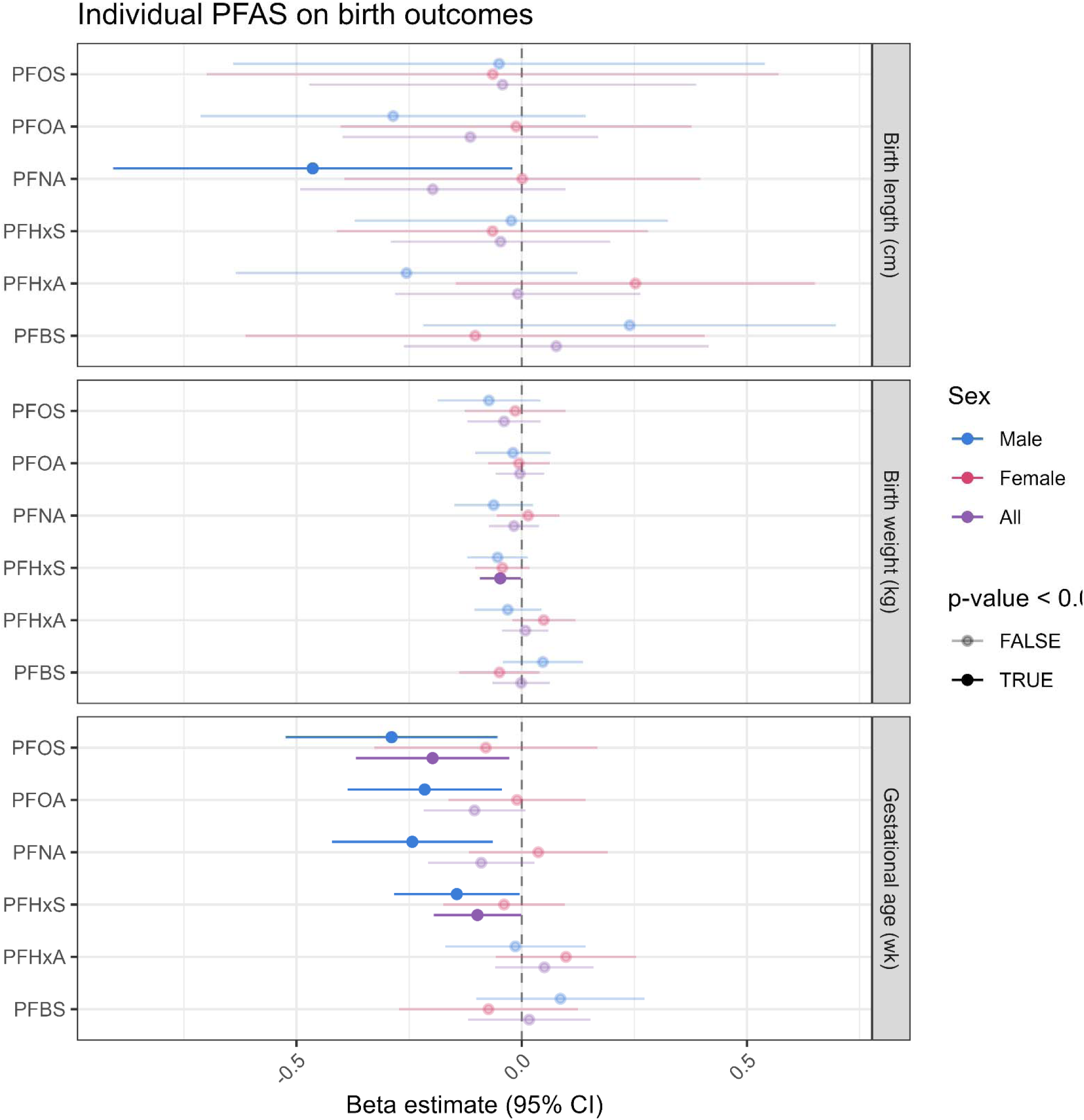
Forest plot depicting prenatal trimester one PFAS-birth outcomes associations. Beta effect estimates correspond to difference in the outcome per doubling in exposure. 95% confidence intervals are depicted. Statistically significant results are fully saturated in their respective colors.

For birth weight, each doubling of PFHxS was associated with lower birth weight in the whole cohort (β = −0.048 kg, 95% CI: −0.093 to −0.002, p = 0.042). For birth length, each doubling of PFNA was associated with shorter length in males (β = −0.464 cm, 95% CI: −0.907 to −0.021, p = 0.042). All single-PFAS results with gestational age, birth weight, and birth length are reported in **Supplemental Table 1a**. The joint effect of the six-PFAS mixture was not significantly associated with gestational age, birth weight, or birth length in the whole cohort or in sex-stratified models (**Supplemental Table 1b**). Examination of the component weights from the mixture fits indicates that this null joint effect reflects statistically offsetting positive and negative congener-specific contributions (**Supplemental Table 1c**). For example, for birth weight in the whole cohort, PFOA constituted the entire positive partition (weight = 1.00, ψ+ = 0.07 kg), which was offset by a negative partition comprising PFHxS (0.33), PFNA (0.26), PFBS (0.17), PFHxA (0.14), and PFOS (0.10; ψ− = −0.15 kg). For gestational age (ψ+ = 0.01 wk versus ψ− = −0.21 wk), the negative weights were dominated by PFOS (0.40) and PFNA (0.39), both of which were inversely associated with gestational age in the single PFAS congener models. For birth length, PFNA carried most of the negative weight (0.79) against PFOA in the positive partition (0.81). Because the two partitions were of comparable magnitude and opposite sign, they largely cancelled within the overall mixture effects. Weight loadings for all birth outcomes for the whole cohort analysis are shown in **Supplemental Figure S1**.

Associations with birth outcomes were largely robust to additional adjustment for maternal hemodynamic markers, serum albumin (g/dL), estimated glomerular filtration rate (eGFR), or both (**Supplemental Figure S2**, **Supplemental Table 1a-c**). All four male-specific gestational-age associations were consistent and remained significant across every adjustment model. Among whole cohort associations, the PFOS association with gestational age was modestly attenuated after eGFR adjustment (p = 0.096), as were the PFHxS associations with gestational age (p = 0.053, 0.12, and 0.11 for albumin-, eGFR-, and jointly adjusted models) and birth weight (p = 0.053, 0.098, and 0.12). Overall, effect estimates were very similar across primary and sensitivity analyses, indicating that loss of precision, rather than hemodynamic confounding, is the more likely explanation. Joint mixture effects remained null for all three birth outcomes under every covariate set.

### Associations between prenatal PFAS and longitudinal growth trajectory parameters

Individual growth trajectories from 0.5 to 24 months were summarized using SITAR size (a) and intensity (c) parameters for weight, length, fat mass, and fat-free mass, and were modeled against each PFAS in single-congener PFAS models adjusted for maternal BMI, age, education, and breastfeeding duration. Higher PFHxS was associated with a smaller growth size across several trajectories. In the whole cohort, each doubling of PFHxS was associated with a smaller length size parameter (β = −0.201 cm, 95% CI: −0.397 to −0.004, p = 0.047, **Figure 2**) and a smaller fat-free mass size parameter (β = −0.047 kg, 95% CI: −0.089 to −0.005, p = 0.031); both associations were stronger in females (length size β = −0.300 cm, 95% CI: −0.592 to −0.009, p = 0.046; fat-free mass size β = −0.077 kg, 95% CI: −0.137 to −0.016, p = 0.015).

**Figure 2.**
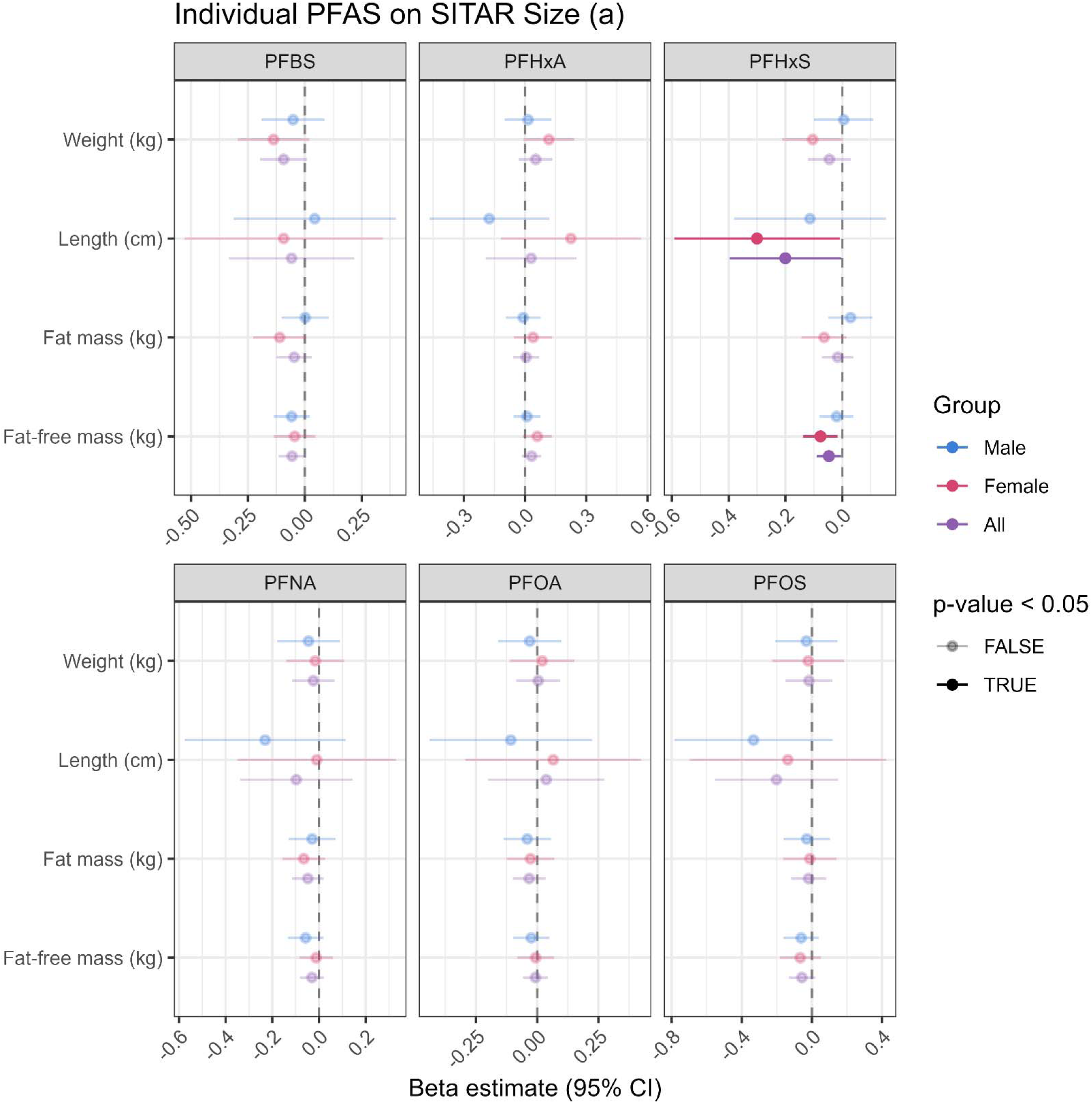
Forest plot depicting prenatal trimester one PFAS-longitudinal growth outcomes associations summarized by the SITAR size (a) parameter, the shift in absolute magnitude of growth trajectory. Beta effect estimates correspond to difference in the outcome per doubling in exposure. 95% confidence intervals are depicted. Statistically significant results are fully saturated in their respective colors.

For the growth intensity (c) parameter, which is dimensionless and is reported here as the percentage difference in growth rate per doubling of exposure, higher PFBS was associated with a lower weight intensity parameter (−1.59%, 95% CI: −2.86% to −0.30%, p = 0.020, Figure 3) and a lower fat-free mass intensity parameter (−1.59%, 95% CI: −2.86% to −0.20%, p = 0.024) in the whole cohort. In females, higher PFHxS was associated with a lower fat-free mass intensity parameter (−1.49%, 95% CI: −2.86% to −0.10%, p = 0.037), whereas higher PFHxA was associated with a greater weight intensity parameter (+2.33%, 95% CI: +0.40% to +4.19%, p = 0.017). All single-PFAS associations with SITAR size and intensity parameters are reported in **Supplemental Table 2a**.

**Figure 3.**
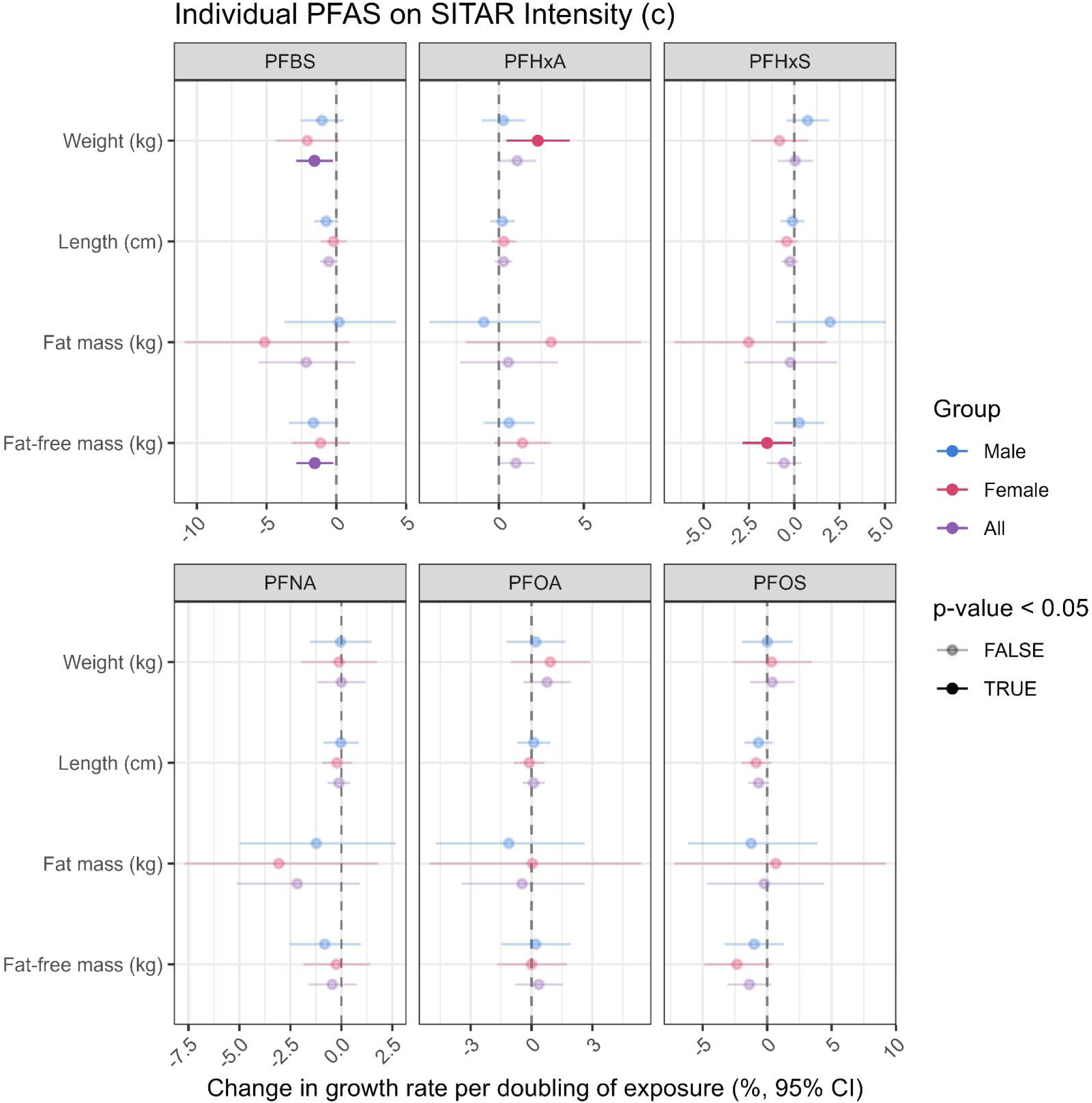
Forest plot depicting prenatal trimester one PFAS-longitudinal growth outcomes associations summarized by the SITAR intensity (c) parameter, the proportional difference in the pace of growth. Estimates correspond to the percentage difference in growth rate per doubling in exposure. 95% confidence intervals are depicted. Statistically significant results are fully saturated in their respective colors.

The joint PFAS mixture was not associated with the size or intensity of infant weight, length, fat mass, or fat-free mass growth during the first two years of life, in the whole cohort or in sex-stratified models (**Supplemental Table 2b**), although a negative trend was observed between the PFAS mixture and fat-free mass size in the whole cohort (Ψ = −0.111 kg, 95% CI: −0.228 to 0.006, p = 0.064). As with the birth outcomes, the component weights from the primary mixture fits indicate that opposing congener-specific contributions offset one another (**Supplemental Table 2c**). For the length size parameter in the whole cohort, the negative partition (ψ− = −0.52 cm) was led by PFHxS (0.55) and PFNA (0.28), while PFOA carried most of the positive partition (0.78; ψ+ = 0.32 cm). For the fat-free mass size parameter, the negative partition (ψ− = −0.15 kg) was distributed across PFBS (0.37), PFOS (0.29), and PFHxS (0.27), against a positive partition of PFOA (0.66) and PFHxA (0.34; ψ+ = 0.04 kg). The weight loadings for weight intensity recapitulated the single-congener results directly: PFBS (0.46) and PFNA (0.39) dominated the negative partition, whereas PFHxA (0.58) and PFOA (0.39) dominated the positive partition. Weight loadings for all SITAR parameters in the whole cohort are shown in **Supplemental Figure S3**.

Overall, additional adjustment for maternal albumin, eGFR, or both yielded similar associations for the growth trajectory analyses (**Supplemental Figure S4-5**, **Supplemental Table 2d-e**). The primary associations between PFBS and lower fat-free mass intensity parameter in the whole cohort, and for PFHxS and smaller fat-free mass size parameter among females remained robust across adjustment models. The positive association between PFHxA and greater weight intensity in females strengthened after adjustment (+3.0% to +3.2%, p = 0.002 to 0.003 depending on adjustment). Several associations were attenuated to non-significance despite nearly identical magnitudes of effect, again likely reflecting reduced precision rather than hemodynamic confounding. A small number of associations reached nominal significance only in the hemodynamically adjusted models, and only for the intensity parameter, including positive associations between weight intensity and PFOA in females, weight intensity and PFOA and PFHxA in the whole cohort, and between PFHxA and fat-free mass intensity in females and the whole cohort. The negative trend for the joint mixture effect on fat-free mass size in the whole cohort was similar in magnitude but no longer trend-level after these additional adjustments (Ψ = −0.102 to −0.091 kg, p = 0.11 to 0.16). No mixture effect reached statistical significance in any model with hemodynamic adjustment.

## Discussion

In this prospective cohort of mother-child dyads from the Central Arkansas region, we examined associations between first trimester maternal PFAS exposure with birth outcomes and detailed infant growth trajectories across the first two years of life. Consistent with the early perinatal growth restriction component of our hypothesis, higher prenatal PFAS concentrations were typically associated with shorter gestation and reduced birth weight and length; we also observed sex-specific differences in postnatal growth trajectories. Reductions in gestational age were most consistent among males and reductions in postnatal size were most consistent among females, while reductions in growth intensity were seen in the whole cohort. For example, PFNA was inversely associated with birth length only among males. The individual PFAS most consistently implicated was PFHxS, which was associated with shorter gestation, lower birth weight, smaller attained length and fat-free mass, and lower female fat-free mass growth intensity, followed by PFBS, which was associated with lower weight and fat-free mass growth intensity in the whole cohort. PFHxA was uniquely associated with greater female weight growth intensity but not size. The joint six-PFAS mixture was not associated with any outcome, but examination of the directional weight loadings suggests that this overall null mixture effect reflects offsetting congener-specific contributions, with PFOA most commonly anchoring the positive partition across nearly every outcome and PFHxS, PFNA, PFBS, and PFOS anchoring the negative partition. Fat mass was not associated with PFAS in either direction, and any increase in later life adiposity may be preceded or masked by the reduced overall growth we observed, particularly among females.

Detectable serum concentrations of PFAS were present in all participants, with PFOS most abundant and the short-chain PFBS least abundant. As previously reported for this cohort, maternal PFOS and PFHxS concentrations were comparable to U.S. NHANES 2011–2014 levels^38^, whereas PFNA and PFOA were somewhat elevated, and overall levels were slightly below the most recent pre-pandemic NHANES 2017-March 2020 estimates^39^. The Glowing cohort is relatively healthy, well-educated, and predominantly White, a profile with comparatively few competing risk factors, which strengthens internal validity even as it limits generalizability.

We focused on first trimester maternal serum PFAS measures because early pregnancy is most frequently characterized in the birth cohort literature, and prior work in our cohort indicates that early gestation samples have the highest PFAS detection rates and are among the most robust sentinels of placental and cord plasma PFAS burden^40^. Additionally, pregnancy-related plasma volume expansion and increased glomerular filtration rate^35,36^ can influence measured PFAS concentrations since PFAS bind to albumin and are cleared renally^34^. Albumin and eGFR have also been associated with anthropometric birth outcomes^41,42^, and some studies have observed confounding by glomerular filtration rate explaining a substantial proportion of a PFOA-birth weight association that increased in later gestation^43–45^. Although hemodynamic changes are less pronounced during early pregnancy, we nevertheless evaluated potential confounding from hemodynamics.

We directly addressed potential confounding by maternal hemodynamics with sensitivity analyses that accounted for first trimester serum albumin, estimated glomerular filtration rate, and both markers simultaneously as covariates and our findings were largely unchanged. The effect estimates for primary birth and growth associations remained highly consistent across adjustment models, while some associations lost nominal significance. Since approximately 15% of the sample did not have hemodynamic measures, this pattern is more consistent with reduced precision rather than with hemodynamic confounding. This mirrors findings in Project Viva, where early-pregnancy associations with birth outcomes were unchanged after adjustment for glomerular filtration and plasma albumin^46^, and in a Spanish cohort where adjustment for eGFR did not alter PFAS-birth weight associations^47^. A small number of associations reached nominal significance only after hemodynamic adjustment but were directionally consistent and similar in magnitude to primary models. Together, these observations indicate that hemodynamic confounding is an unlikely explanation of the associations reported here.

Our findings of shorter gestational age predominantly in males for multiple PFAS, lower birth weight associated with PFHxS overall, and shorter birth length with PFNA in males, align with a body of evidence linking prenatal PFAS exposure to shorter gestation, preterm birth, and smaller birth size, particularly for longer-chain compounds^6,21,48,49^, although some cohorts report null associations^50^. However, other cohorts have also reported PFAS-associated smaller birth size that was strongest among females^21^. Our male-predominant findings comport with the Clifton hypothesis, that male placentae may prioritize in utero growth over stress adaptation, potentially making them more vulnerable to adverse birth outcomes^23^. Prior reports have also identified PFAS-related male susceptibilities. First trimester PFOS and PFHxS have been associated with gestational hypertension in male but not female pregnancies, although a PFHxS-preeclampsia association was found only among female fetuses^51^. Another study identified an association between placental PFDA and shorter gestational age only in males^52^. Our group identified an association between a placental PFAS mixture and decreased estimated syncytiotrophoblast proportions at term among male fetuses in a subset of this cohort^53^ in addition to placental DNA methylation differences^40^. Additionally, while the six-PFAS mixture was not associated with any birth outcome, a negative trend for gestational age in males paralleled the single congener PFAS results. Others have similarly reported small PFAS-mixture associated reductions in birthweight that failed to achieve statistical significance^7,54,55^. Our analysis showed that directional weight loadings reflected offsetting congener-specific contributions, indicating that future work should carefully consider mixture operationalization alongside PFAS-specific effects.

For postnatal growth trajectory analyses, PFHxS was associated with smaller size for length and fat-free mass and with lower female fat-free mass growth intensity and PFBS with lower weight and fat-free mass growth intensity in the whole cohort whereas PFHxA was associated with greater female weight intensity. Because the SITAR intensity parameter indexes the rate of growth, this reflects a higher peak growth velocity for PFHxA rather than a departure from the broader pattern, and it is consistent with PFHxA loading positively in the mixture analysis weights. Overall, individual PFAS were predominantly associated with smaller and slower infant growth, concentrated among females. This inverse, female-predominant direction is supported by several comparable longitudinal cohorts. In the Healthy Start study, PFOS and PFHxS were associated with lower weight-for-age z-scores among female infants at approximately five months of age^56^. A New York State cohort reported smaller growth trajectories from birth to 3 years with higher neonatal PFOS and PFOA measured from newborn dried blood spots among girls only^57^, and a Taiwanese cohort reported decreased weight and BMI z-scores with cord PFOS among girls specifically across the 6–12 and 12–24 month intervals^58^. These results are consistent with the broader synthesis that, within the first two years of life, prenatal PFAS exposure shows no clear association with length and inconsistent negative associations with weight and BMI, in contrast to positive adiposity associations that emerge in later childhood and adolescence^49,58^. Those later-childhood, female-predominant increases have been documented with directly measured body composition including higher DXA fat mass index among girls in mid-childhood in Project Viva^15^ and higher DXA total, gynoid, and android fat among girls at age 7 in the Odense Child Cohort, with null associations in boys^59^.

Not all comparable evidence aligns with the patterns we observed. In the New Hampshire Birth Cohort Study, growth mixture modeling of infant BMI across the first year found that girls born to mothers with higher pregnancy PFOS concentrations were more likely to follow higher-velocity growth trajectories, a female-centric pattern opposite in direction to ours^60^. In the SELMA cohort, a shape-invariant model up to age 5.5 years linked higher prenatal PFOA to a later age at peak growth velocity but also to heavier infant weight at the weight plateau among girls^61^. In the Healthy Start study, maternal PFOA and PFNA were positively associated with percent fat mass at 5 months among male infants, while our findings do align with their observed associations for PFOS and PFHxS among females^56^. There are several important methodological differences that may explain these distinctions. The contradicting results focused on BMI or percent fat mass, whereas our female-predominant associations were primarily for fat-free mass, for which fewer comparable data exist. Because BMI and other corrective indices such as fat-free mass index were poorly statistically modelled by SITAR in our dataset with preliminary analyses in this age range, we did not investigate them here. PFOS was not among the congeners associated with SITAR parameters in our cohort and exposure contrasts differ across studies. We note also that one of our findings, PFHxA associated with greater female weight intensity, is directionally concordant with the positive female associations reported in SELMA^61^ and with PFHxA’s consistently positive weight loading in our mixture results. Mechanistically, sex-specific effects are consistent with the recognized endocrine-disrupting activity of PFAS, including activation of peroxisome proliferator-activated receptors and alteration of steroid hormone synthesis, together with weak estrogenic or anti-androgenic properties^49^, and evidence that prenatal PFAS may alter testosterone concentrations in female offspring^62^.

The joint PFAS mixture was not associated with any birth or growth parameter, with only a negative trend for the fat-free mass size parameter in the whole cohort. Examination of the directional weight loadings from the primary mixture models revealed consistent off-setting contributions. Across nearly every outcome, PFOA anchored the positive partition while PFHxS, PFNA, PFBS, and PFOS anchored the negative partition. This pattern recapitulated single congener model results. Consequently, a simultaneous one-quartile increase in all six congeners may not be the most informative exposure contrast when components act in opposition, since a summary mixture effect can be null while a coherent directional subset is present. However, positive and negative weights are descriptive quantities reported without confidence intervals or hypothesis tests when directional homogeneity fails^63^. Generally, testing exposure contrasts reflecting actual environmental source-mixtures or a priori chemical classes may translate estimated health effects more directly into interventions on the real-world sources that generate them^63,64^.

Several limitations should be considered. The analytic sample of 217 dyads, and particularly the sex-stratified subsets (93 females), provided limited statistical power to test apparent sex-specific associations. Exposures were measured in 2010–2014 and may not reflect current PFAS exposures, and the cohort’s predominantly White, well-educated composition, and exclusion of miscarriages, stillbirths, and gestational diabetes cases, while advantageous for internal validity, may limit external generalizability. Creatinine-based estimating equations for glomerular filtration were derived in non-pregnant populations and may perform poorly in pregnancy^42,65^, though we also considered serum albumin as an alternative. The SITAR timing parameter could not be modeled owing to convergence limitations, constraining our characterization of growth tempo. Our study also has several strengths. Direct assessment of adiposity and systematic evaluation of hemodynamic markers address concerns that have been raised repeatedly but have been assessed directly in few cohorts^46^. Nonlinear and shape-invariant growth models have previously been applied to infant peak growth velocity and its timing in relation to prenatal PFOA^61^ and an endocrine disrupting chemical mixture to a slower infant growth spurt^48^. However, our application to infant fat and fat-free mass, and testing their association with PFAS mixtures, is to our knowledge novel, as is this examination of prenatal PFAS and infant growth in the Central Arkansas region.

In this Central Arkansas cohort, first trimester maternal PFAS concentrations were individually associated with modestly shorter gestation and with reduced early growth, length, and fat-free mass, with sex-specific patterns in which males were more affected at birth and females during postnatal growth. These associations were small but directionally consistent with known PFAS toxicity, were detectable in a relatively healthy, low-risk population, and were robust to adjustment for maternal hemodynamic markers. Fat mass was not associated with PFAS during the first two years of life, suggesting that early PFAS-related alterations in body composition may not be reflective of early adiposity, but instead of reduced lean tissue accrual. Longer-term follow-up will be important to determine whether these early sex-specific growth patterns persist or lead to later life differences in adiposity and other growth outcomes.

## Data availability statement

Data are available upon reasonable request contingent on data privacy and access regulations.

## Acknowledgements

We thank the participants who provided biospecimens for this study. This work was supported by funding from the National Institute of Environmental Health Sciences, NIH/NIEHS [R01 ES032176; MPIs: Andres, Everson, & Pearson], NIH/NIEHS [R01 ES036986; MPIs: Andres & Everson], the HERCULES Center [P30 ES019776; MPIs: Everson, Barr], the UK-CARES Center [P30 ES026529; CoI: Pearson]. KS is supported by USDA ARS CRIS 3093-10700-001-000-D. AJM is a Veterans Affairs research career scientist and this work used resources at the Central Arkansas Veterans Affairs Healthcare System. The content is solely the responsibility of the authors and does not necessarily represent the official views of the National Institutes of Health. This manuscript is the result of funding in whole or in part by the National Institutes of Health (NIH). It is subject to the NIH Public Access Policy. Through acceptance of this federal funding, NIH has been given a right to make this manuscript publicly available in PubMed Central upon the Official Date of Publication, as defined by NIH.

Generative AI tools were used in the preparation of this work. Claude Opus and Sonnet model families (*Anthropic*, San Francisco, CA) were used to assist in drafting and reviewing the R analysis code, to draft and critically review sections of the manuscript, and to assist in preparing the Table of Contents graphic. All AI-generated code was reviewed, tested, and verified against the analytic dataset by the authors; all AI-drafted text was reviewed, rewritten, and edited by the authors; and all AI-assisted suggestions in text or code were independently evaluated before adoption. No participant-level data were provided to any AI tool. The authors take full responsibility for the content, accuracy, and integrity of this manuscript.

## Author contributions statement

**Kyle A. Campbell:** Conceptualization, Formal analysis, Methodology, Investigation, Data Curation, Writing – Original Draft, Software, Validation, Visualization

**Clark R. Sims**: Conceptualization, Formal analysis, Methodology, Investigation, Data Curation, Writing – Original Draft, Writing – review & editing

**Priyadarshni Patel:** Formal analysis, Methodology, Data Curation, Writing – review & editing

**Lauren A. Havens:** Data curation, Investigation, Writing – review & editing

**Andrew J. Morris:** Data curation, Funding acquisition, Investigation, Project administration, Resources, Supervision, Validation, Writing – review & editing

**Kartik Shankar:** Conceptualization, Data curation, Funding acquisition, Investigation, Project administration, Resources, Validation, Writing – review & editing

**Kevin J. Pearson:** Funding acquisition, Project administration, Resources, Writing – review & editing

**Aline Andres:** Conceptualization, Data curation, Funding acquisition, Investigation, Methodology, Project administration, Resources, Supervision, Validation, Writing – review & editing

**Todd M. Everson:** Conceptualization, Data curation, Formal analysis, Funding acquisition, Methodology, Project administration, Resources, Software, Supervision, Validation, Writing – original draft, Writing – review & editing

## Abbreviations

4:2 FTS: 4:2 fluorotelomer sulfonic acid
6:2 FTS: 6:2 fluorotelomer sulfonic acid
8:2 FTS: 8:2 fluorotelomer sulfonic acid
9Cl-PF3ONS: 9-chlorohexadecafluoro-3-oxanonane-1-sulfonic acid
11Cl-PF3OUdS: 11-chloroeicosafluoro-3-oxaundecane-1-sulfonic acid
BMI: Body Mass Index
CI: Confidence Interval
DONA: 4,8-dioxa-3H-perfluorononanoic acid
FFM: Fat-free Mass
HFPO-DA: hexafluoropropylene oxide dimer acid
LOD: Limit of Detection
N-EtFOSAA: n-ethyl perfluorooctane sulfonamido acetic acid
NHANES: National Health and Nutrition Examination Survey
N-MeFOSAA: n-methylperfluorooctanesulfonamidoacetic acid
PFAS: Per- and Polyfluoroalkyl Substances
PFBA: perfluorobutanoic acid
PFBS: perfluorobutane sulfonate
PFDA: perfluorodecanoic acid
PFDoA: perfluorododecanoic acid
PFDS: perfluorodecanesulfonic acid
PFHpA: perfluoroheptanoic acid
PFHpS: perfluoroheptanesulfonic acid
PFHxA: perfluorohexanoic acid
PFHxS: perfluorohexane sulfonate
PFNA: perfluorononanoic acid
PFNS: perfluorononanesulfonic acid
PFOA: perfluorooctanoic acid
PFOS: perfluorooctane sulfonic acid
PFOSA: perfluorooctanesulfonamide
PFPeA: perfluoro-n-pentanoic acid
PFPeS: perfluoropentane sulfonic acid
PFTeDA: perfluorotetradecanoic acid
PFTrDA: perfluorotridecanoic acid
PFUnDA: perfluoroundecanoic acid
SITAR: SuperImposition by Translation And Rotation

