## Supplemental Info for "PFAS Exposure During Pregnancy Is Associated with Shorter Gestational Age at Birth and with Growth Trajectories During the First 2 Years of Life"

Author Affiliations:

### Supporting Information for Publication

**Supplemental Table 1.** Associations between first-trimester maternal PFAS exposure and birth outcomes (gestational age, birth weight, birth length) from single-congener PFAS linear regression and joint six-PFAS quantile g-computation models, in the whole cohort and by infant sex, stacked across four covariate-adjustment models (primary; +albumin; +eGFR; +albumin & eGFR). A README sheet defines the covariate sets and every column. Sheet tabs report individual-PFAS associations (S1a), joint mixture effects (S1b), and mixture component weights with their positive and negative partition sums (S1c).

**Supplemental Table 2.** Associations between first-trimester maternal PFAS exposure and SITAR-derived infant growth trajectory parameters (size and intensity, across weight, length, fat mass, and fat-free mass) from single-congener PFAS linear regression and joint six-PFAS quantile g-computation models, in the whole cohort and by infant sex, stacked across four covariate-adjustment models (primary; +albumin; +eGFR; +albumin & eGFR). Size (a) estimates are in outcome units; intensity (c) is dimensionless and is additionally reported as the percentage difference in growth rate (Pct_Change, Pct_CI_95). A README sheet defines the covariate sets and every column. Sheet tabs report individual-PFAS associations (S2a), joint mixture effects (S2b), mixture component weights with their positive and negative partition sums (S2c), and covariate-set comparisons for individual-PFAS (S2d) and mixture (S2e) effects.

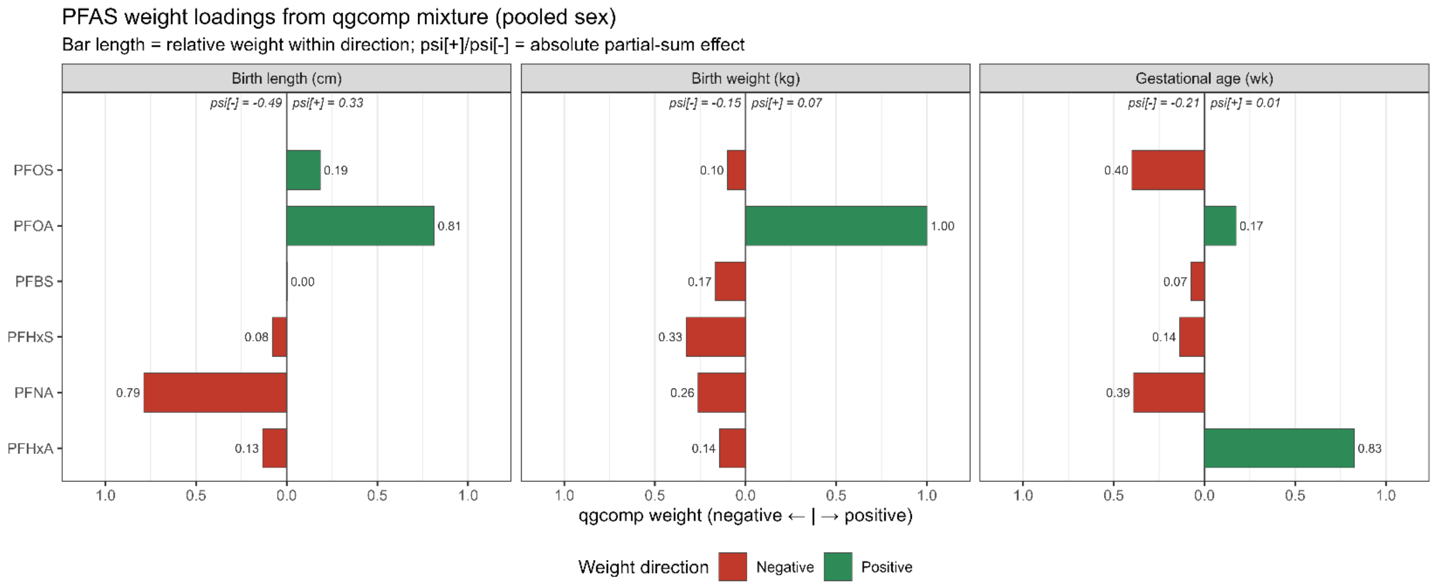


**Supplemental Figure S1.** Summary of the whole cohort quantile g-computation six-PFAS joint mixture testing associations with birth outcomes. Negative and positive partial mixture effects are listed as psi[-] and psi[+], respectively. The relative contribution of each PFAS to the negative or positive partial effects are summarized in green for positive contributions and red for negative contributions. Model results are presented for each birth outcome.

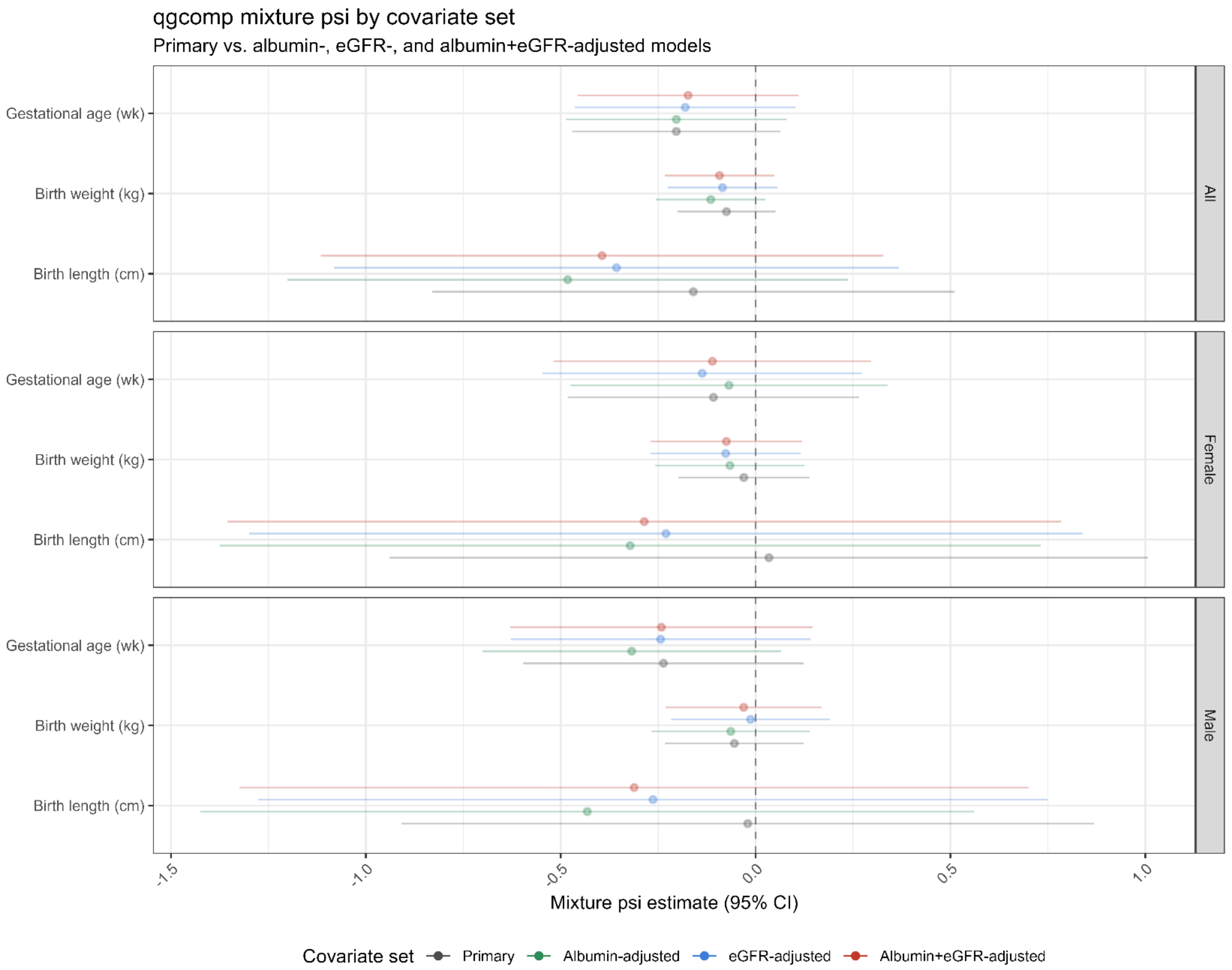


**Supplemental Figure S2.** Hemodynamic sensitivity analysis quantile g-computation model results forest plot for birth outcomes stratified by whole cohort and sex. The primary model presented in grey, albumin-adjusted in green, eGFR-adjusted in blue, and simultaneous adjustment for serum albumin and eGFR in red. Results significant at alpha = 0.05 highlighted in full color.

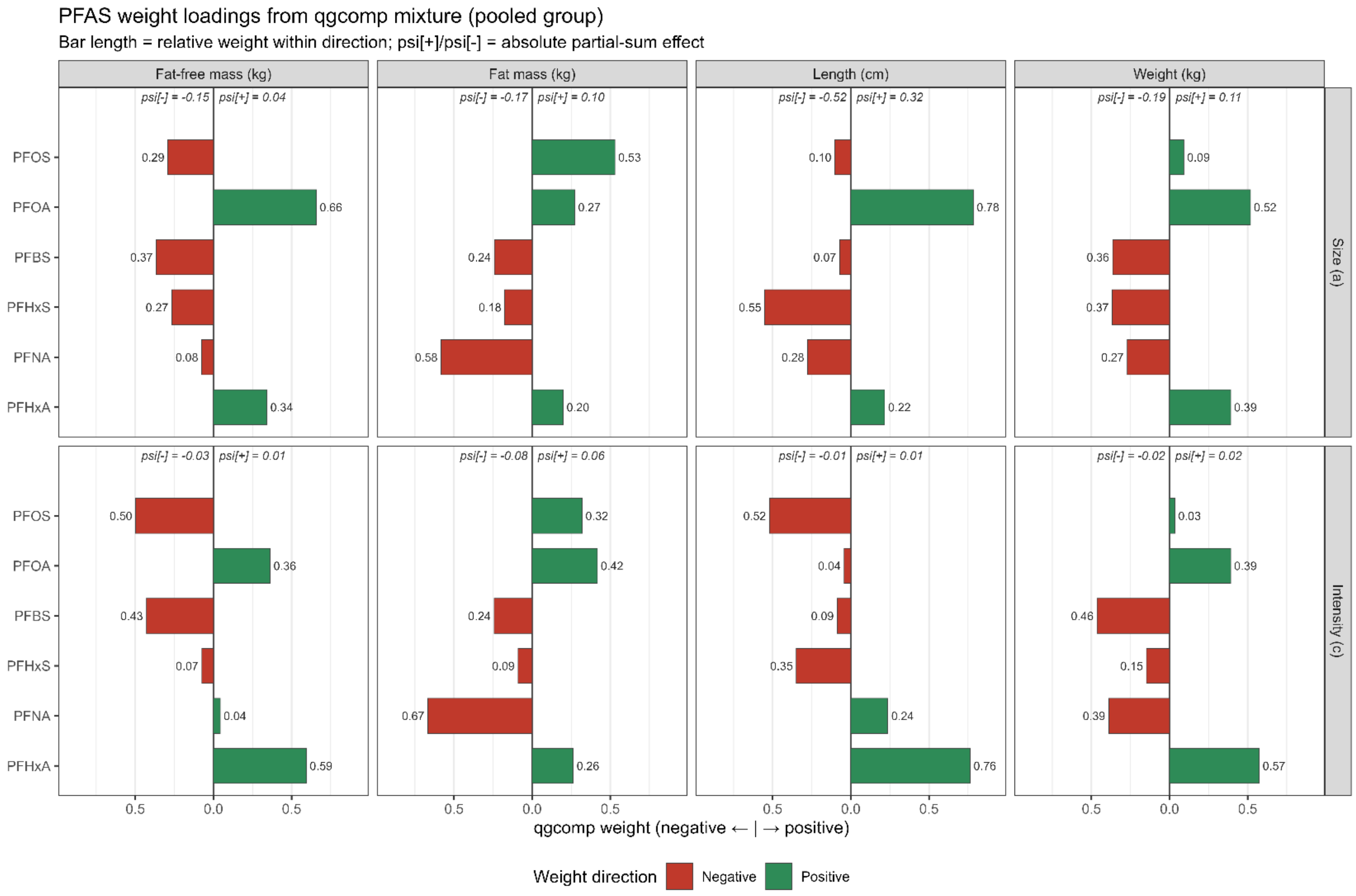


**Supplemental Figure S3.** Summary of the whole cohort quantile g-computation six-PFAS joint mixture testing associations with postnatal growth trajectory parameters. Negative and positive partial mixture effects are listed as psi[-] and psi[+], respectively. The relative contribution of each PFAS to the negative or positive partial effects are summarized in green for positive contributions and red for negative contributions. Model results are presented for each growth parameter outcome.

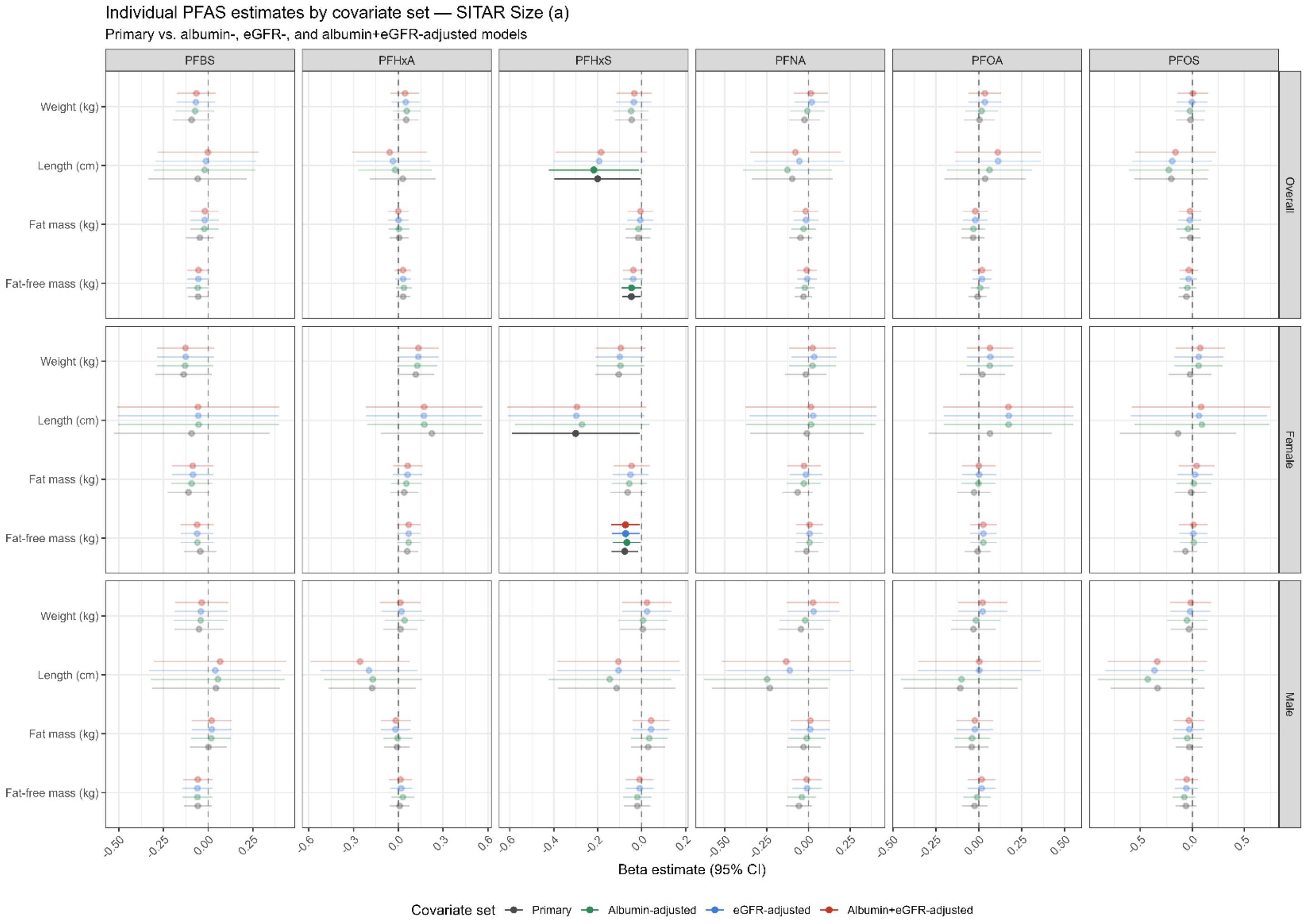


**Supplemental Figure S4.** Hemodynamic sensitivity single-congener PFAS linear regression results forest plot testing the association between infant growth outcomes' SITAR-summarized size (a) parameter and a doubling in an individual PFAS stratified by whole cohort and sex. The primary model presented in grey, albumin-adjusted in green, eGFR-adjusted in blue, and simultaneous adjustment for serum albumin and eGFR in red. Results significant at alpha = 0.05 highlighted in full color.

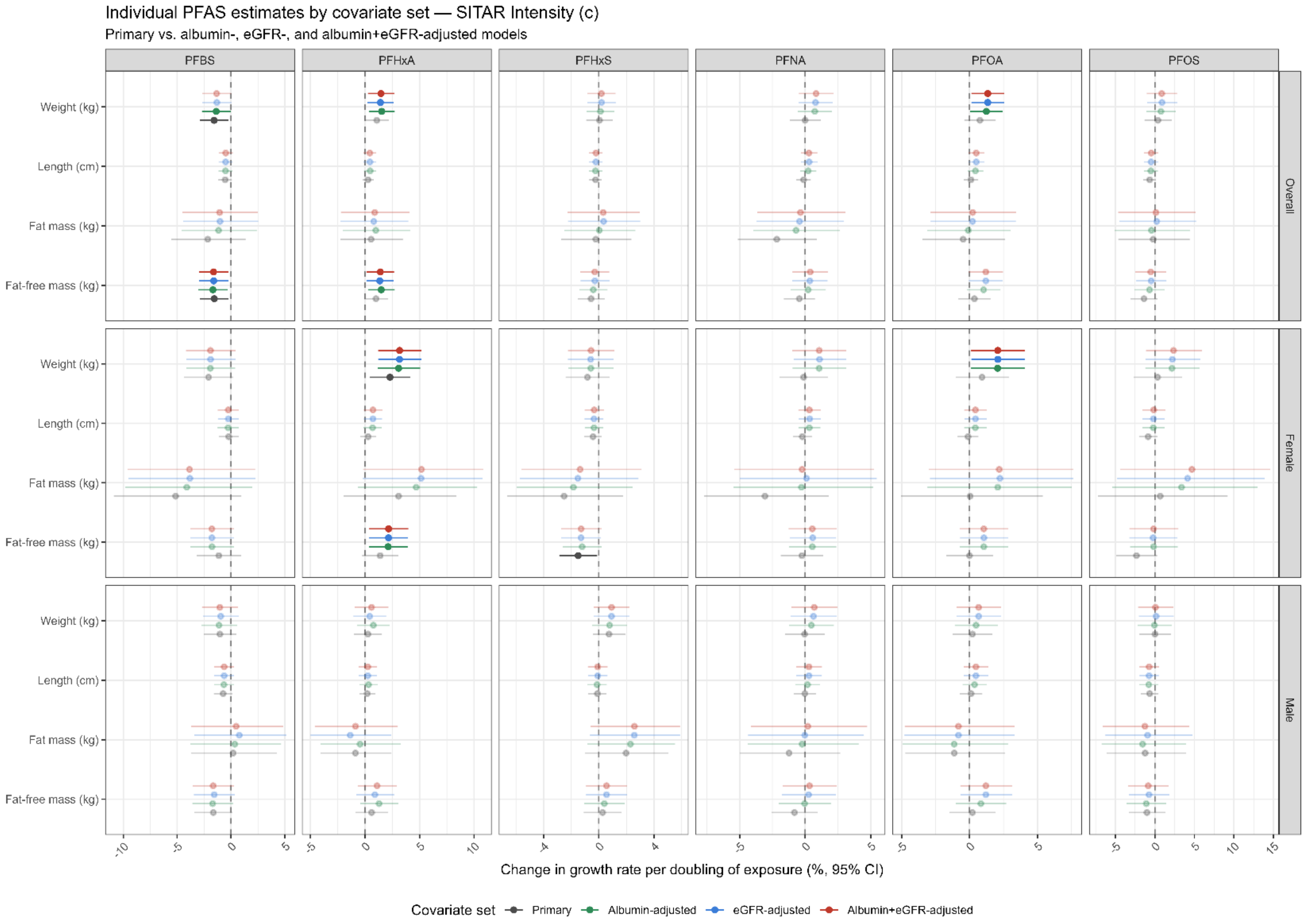


**Supplemental Figure S5.** Hemodynamic sensitivity single-congener PFAS linear regression results forest plot testing the association between infant growth outcomes' SITAR-summarized intensity (c) parameter and a doubling in an individual PFAS stratified by whole cohort and sex. The intensity parameter is dimensionless; estimates correspond to the percentage difference in growth rate per doubling in exposure. The primary model presented in grey, albumin-adjusted in green, eGFR-adjusted in blue, and simultaneous adjustment for serum albumin and eGFR in red. Results significant at alpha = 0.05 highlighted in full color.
